# Effectiveness of Second Wave COVID-19 Response Strategies in Australia

**DOI:** 10.1101/2020.11.16.20232843

**Authors:** George Milne, Simon Xie, Dana Poklepovich, Dan O’Halloran, Matthew Yap, David Whyatt

## Abstract

**Background:** There is a significant challenge in responding to second waves of COVID-19 cases, with governments being hesitant in introducing hard lockdown measures given the resulting economic impact. In addition, rising case numbers reflect an increase in coronavirus transmission some time previously, so timing of response measures is highly important. Australia experienced a second wave from June 2020 onwards, confined to greater Melbourne, with initial social distancing measures failing to reduce rapidly increasing case numbers. We conducted a detailed analysis of this outbreak, together with an evaluation of the effectiveness of alternative response strategies, to provide guidance to countries experiencing second waves of SARS-Cov-2 transmission.

**Method:** An individual-based transmission model was used to 1) describe a second-wave COVID-19 epidemic in Australia; 2) evaluate the impact of lockdown strategies used; and 3) evaluate effectiveness of alternative mitigation strategies. The model was calibrated using daily diagnosed case data prior to lockdown. Specific social distancing interventions were modelled by adjusting person-to-person contacts in mixing locations.

**Results:** Modelling earlier activation of lockdown measures are predicted to reduce total case numbers by more than 50%. Epidemic peaks and duration of the second wave were also shown to reduce. Our results suggest that activating lockdown measures when second-wave case numbers first indicated exponential growth, would have been highly effective in reducing COVID-19 cases. The model was shown to realistically predict the epidemic growth rate under the social distancing measures applied, validating the methods applied.

**Conclusions:** The timing of social distancing activation is shown to be critical to their effectiveness. Data showing exponential rise in cases, doubling every 7-10 days, can be used to trigger early lockdown measures. Such measures are shown to be necessary to reduce daily and total case numbers, and the consequential health burden, so preventing health care facilities being overwhelmed. Early control of second wave resurgence potentially permits strict lockdown measures to be eased earlier.

All authors have seen and approved the manuscript. Research funding from Department of Health, Western Australia and Department of Health, Queensland is acknowledged. The authors confirm that these organisations had no influence on the submitted work, nor are there any competing interests.

## Background

An ongoing challenge faced by public health authorities during the COVID-19 pandemic is knowing when to activate social distancing strategies, the magnitude of the measures, how to safely ease the measures once case numbers reach low levels, and how best to react when a rapidly developing outbreak occurs. We report on a detailed, model-based case study into the significant COVID-19 second wave in greater Melbourne, Australia, from June 2020 onwards. This occurred as a result of ineffective management of the mandated hotel quarantine policy for returning overseas travellers, where transmission is believed to have occurred between infectious arrivals and security staff, then among the staff, and thus to their families and friends [1, 2].

The aim of this study was to understand why this second wave of SARS-Cov-2 transmission grew so rapidly; why the initial increase in social distancing response was ineffective; what responses would have been more effective; and thus the lessons learned. The insights gained are of benefit to other countries and jurisdictions in their determination of response policy.

The Australian response to COVID-19 has kept the country largely free from large-scale transmission, such as occurred in Europe, the USA and Latin America, by halting flights from China in February, and stopping inbound travel by non-Australian residents from 20^th^ March 2020. Robust social distancing measures were also adopted country-wide on that date and further strengthened on 26^th^ March 2020 [3-5], resulting in substantial reductions in person-to-person contact in workplaces and the community. These measures resulted in very low levels of COVID-19 virus transmission, with almost all cases arising from returning Australians infected outside the country. All arrivals were required to enter 14 days of managed hotel quarantine. Certain Australian states (*e*.*g*. Western Australia and Queensland) also closed their interstate borders. The initial success of Australia in halting the arrival of infectious individuals, and managing the limited number of community transmissions via testing and contact tracing, allowed all Australian states to start easing social distancing measures from May onwards, including allowing schools to reopen, as detailed in Table 1. The total number of COVID-19 related deaths for the whole of Australia was ∼100 up to 6^th^ May 2020, and stayed constant at that number for the next 8 weeks. In late May, a breakdown in hotel quarantine regulations in Melbourne, State of Victoria, followed by a number of unauthorised, large-scale family gatherings, allowed the SARS-Cov-2 virus to enter the wider population in greater Melbourne, with diagnosed case numbers increasing from June onwards [6].

**Table 1:** Social distancing measures applied in greater Melbourne [3-5, 24, 25]. WN: percentage workplace non-attendance; SC: percentage reduction in school attendance; CCR: percentage reduction in community-wide contact; CI: percentage adult case isolation, all child cases up to and including age 17 isolate.

| Date measures implemented | Social distancing measures | Estimated social distancing |
| --- | --- | --- |
| 25<sup>th</sup> March | **Stage 2** restrictions commence |  |
| 26<sup>th</sup> May | Pre-primary, years 1, 2, 11, 12 students return to school, after holiday period. | WN20<br>SC30 CCR20<br>CI70 |
| 9<sup>th</sup> June | Years 3 to 10 back in school. | WN20<br>SC0<br>CCR20<br>CI70 |
| 26<sup>th</sup> June | School holidays start (as planned). | WN20<br>SC100<br>CCR20<br>CI70 |
| 2<sup>nd</sup> July | **Stage 3** Stay-at-Home restrictions in ten Melbourne postcode areas. | WN20<br>SC100 CCR60<br>CI70 |
| 10<sup>th</sup> July | **Stage 3** Stay-at-Home restrictions extended to all of greater Melbourne. | WN20<br>SC100<br>CCR60<br>CI70 |
| 20<sup>th</sup> July | All ages return to school. | WN20<br>SC0<br>CCR60<br>CI70 |
| 3<sup>rd</sup> August | **Stage 4** restrictions applied to greater Melbourne, including 8pm to 5am curfew. All schools closed. | WN50<br>SC100<br>CCR80<br>CI70 |

Case data resulting from this second wave outbreak provided us with a unique opportunity to analyse the non-pharmaceutical (NPI) measures used, and the significance of their activation timing. This study was facilitated by the fact that Australia’s second wave was geographically contained to greater Melbourne, and not impacted by the ongoing introduction of infectious persons. This “self-contained” second wave provided us with high quality data on daily case numbers, as in Figure 1, allowing us to evaluate second wave response measures without interference from introduced cases.

**Figure 1:**
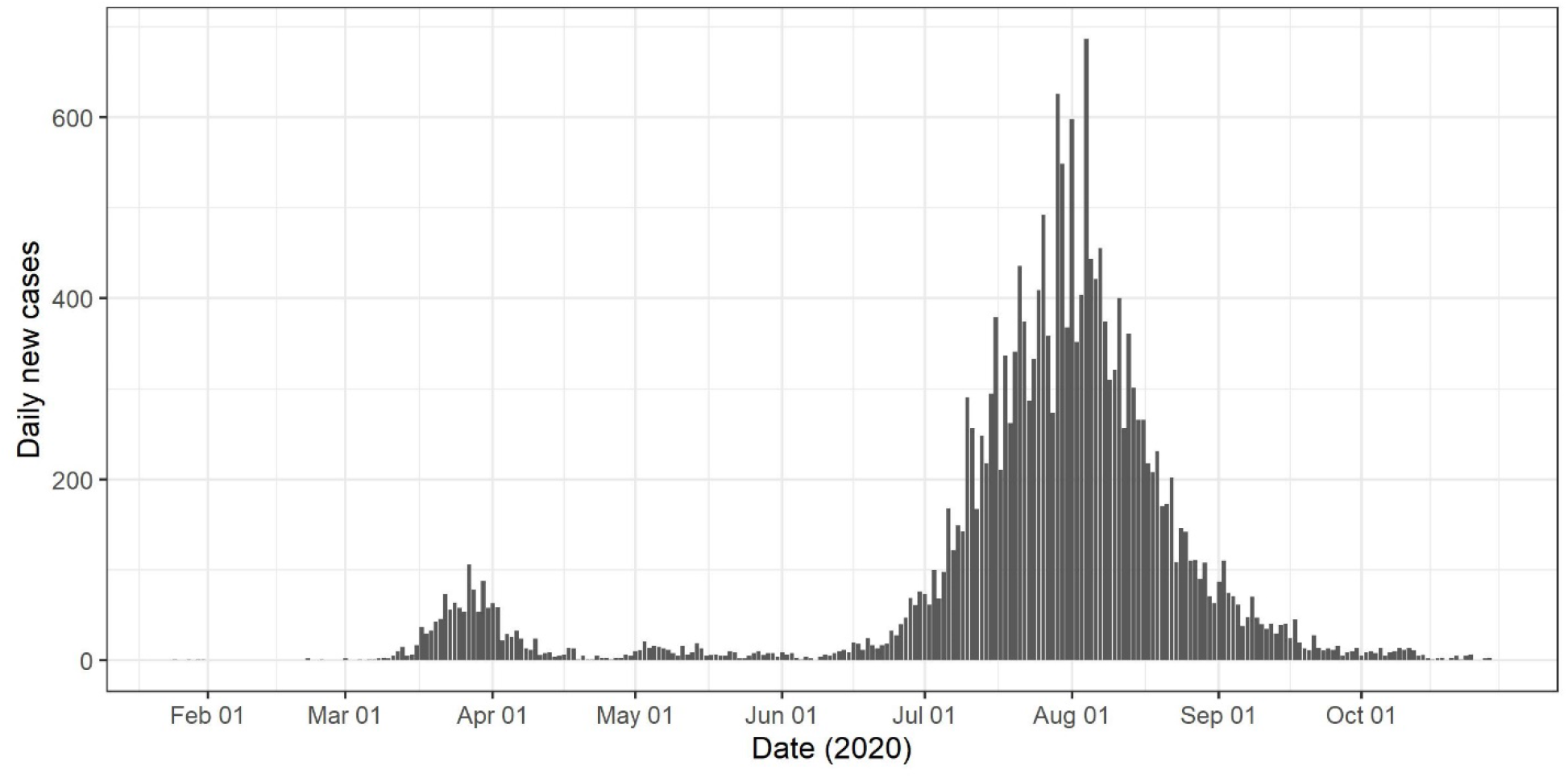
Daily COVID-19 case numbers State of Victoria, Australia. Total of 19,000 cases in greater Melbourne, and 1,000 in regional Victoria, to 30^th^ October 2020 [7, 8].

## Methods

An individual-based model capturing the demographics and movement patterns of individuals within an Australian city, together with SARS-CoV-2 virus transmission data from the early outbreak in Wuhan, China prior to social distancing activation [9], was developed and applied. This was used to analyse the effectiveness of a broad suite of non-pharmaceutical, social distancing interventions, by varying their strength, their time of activation, and their duration. Individual-based (c.*f*. agent-based) modelling is an appropriate methodology to adopt for this task. It permits the effect of four key social distancing measures to be readily captured at a high degree of detail: school closure; reduction in workplace participation; community-contact reduction; and case isolation. This modelling method has been applied previously, to quantify the impact of pandemic mitigation strategies [10-13], and to inform policy decision making [14-17].

This study utilised a model of Newcastle, a city in New South Wales, Australia (population 272,407), whose population demographics reflect Australia as a whole and results were scaled to greater Melbourne, population ∼5 million, following an approach used previously [11, 12]. Australian Bureau of Statistics (ABS) census data were used to capture age-specific demographics of every household in the community.[18, 19] ABS workplace data was used to assign adults to workplaces [20], and State Government schools data was used to assign children to age-specific classes [21]. These data were used to model the time-changing contact patterns for each individual, as they move between their household, school/workplace contact hub and in the wider community.

Model parameter settings were calibrated to reflect the transmission characteristics of the COVID-19 epidemic: an incubation period averaging 6 days, from infection to symptom emergence (if any); a latent period averaging 5 days, from infection to infectious; an infectious period averaging 4.5 days, the first day being asymptomatic; and 35% of cases are asymptomatic. The probability of virus transmission from infectious to susceptible individuals was derived from a R0 of 2.25, based on SARS-CoV-2 transmission characteristics from Wuhan, China prior to introduction of containment measures [9, 22], following the method applied for pandemic influenza [23].

Model outputs obtained by running the simulation software produced the infection history of every individual in the community, generating the daily (and total) number of infectious individuals, and determining where and when infection occur, as described previously [14, 17]. Modelling analyses were conducted for alternative social distancing strategies, by varying the strength of measures and their activation timing. This quantified how alternative mitigation strategies may have performed, allowing us to contrast alternative mitigation strategies with those that were used. The difference in total case numbers provides a measure of the effectiveness of alternative mitigation strategies to reduce the impact of the outbreak.

Our model depends on a small number of stochastic parameters, including the probability of virus transmission between an infectious individual and a susceptible individual, and the random seeding of infectious individuals into the community to initiate an outbreak. This results in variation between successive simulation runs. From experience with prior, related analyses of mitigation strategy effectiveness for pandemic and seasonal influenza [14, 15, 23], averaging infection data generated from multiple runs stabilises after approximately 16 runs. The results presented here were obtained from multiple simulation runs for each social distancing scenario evaluated.

### Social distancing

Four social distancing measures are available to health authorities, and were combined during the Melbourne COVID-19 outbreak. School closure (SC): reduction in school attendance. Workplace non-attendance (WN): a percentage of all persons in the workforce remain at home during working hours. Community contact reduction (CCR): contact in the wider community is reduced by a given percentage to reflect strength of intervention. Case isolation (CI): a percentage of adults and all children withdraw to the home on becoming symptomatic.

Stage 2, Stage 3 and Stage 4 measures applied in greater Melbourne are described in Table 1 and the Supporting Information [3-5, 24, 25]. These have increasing strength, with Stage 4 lockdown restricting individuals to their homes unless they have approved occupations, require healthcare, or require to shop for essential supplies. The sequence of changes to social distancing measures in the State of Victoria are as follows: Stage 2 measures were activated on 25^th^ March, and given absence of community-wide transmission started to be eased on 26^th^ May, with some students returning to schools. From 26^th^ June all schools closed, mirroring Stage 2 measures. Stage 3 measures were introduced from 2^nd^ July onwards, first to limited areas of greater Melbourne, designated by their postal codes. Stage 3 measures were then extended to all of greater Melbourne on 10^th^ July, however schools reopened on 13^th^ July for years 11 and 12, and for all ages on 20^th^ July. As daily case numbers continued to increase over that period, Stage 4 lockdown measures were activated on 3rd August.

### Model calibration

The “second wave” Melbourne COVID-19 outbreak originated in late May 2020, as a result of a breakdown in hotel quarantine [1]. The scale of the outbreak became apparent later, as diagnosed case numbers started to increase rapidly. A calibration process was used to estimate the initial phase of the outbreak, to create a sufficient pool of infectious individuals that later transition into diagnosed cases, and thus align with daily reported case numbers. The calibration aligned the daily number of infectious individuals in the population generated by the simulation model, to actual, reported case numbers. This allowed us to predict case number dynamics into the future, and under alternative social distancing measures.

Calibrating diagnosed cases to new SARS-Cov-2 infections in the simulation model used a process which applied the infection timeline parameters (see Supporting Information, Table S1). The daily set of currently infectious individuals generated by the model was used to calculate the daily number of (simulated) diagnosed cases, assuming a given percentage of COVID-19 positive cases are tested and reported. By adjusting seeding of infectious individuals into the model in early June 2020, and under the social distancing measures then in place, the model was calibrated to case numbers appearing at a later time.

The calibration process was conducted in two steps, to reflect an increased testing regime beginning in early July. The first calibration phase resulted in approximately a third of all infected individuals being diagnosed, the remaining two thirds being in either the latent period, or asymptomatic, or exhibiting mild symptoms, and thus not accessing a testing facility. In the second phase, when a more aggressive testing regime involving increased contact tracing was initiated from 2^nd^ July 2020 onwards, we assumed a steady increase in the (positive case) diagnosis rate, as the overall testing coverage increased. This ratio, between infections and the percentage of infections being diagnosed and reported as cases, was used to convert daily infection data generated by the model into predicted case numbers.

### Strengthened social distancing

Simulation experiments were conducted by running model software, after adjusting the strength of social distancing measures to reflect introduction of Stage3 and Stage 4 restrictions. Random seeding of infectious individuals into the model was used to capture the effect of localized high transmission events, resulting from a number of large gatherings in late June 2020 [2]. The following were determined to replicate daily outbreak case dynamics up to 1^st^ August 2020.

Prior to increases in social distancing measures on 9^th^ July 2020, we assumed 30% of school-age children were not attending school (SC30) and workplace attendance and community contact were both reduced by 20% (WN20) and (CCR20) respectively, as in Table 1. This weak Stage 2 social distancing arose from previous easing of measures, given low levels of virus transmission throughout Australia. Simulations were run from 17^th^ June 2020 onwards with approximately 10 infections seeded daily, and with social distancing at (SC30+WN20+CCR20), as above. From 28^th^ June an additional 180 infected individual were seeded daily to model the rapid growth in infections due to large-scale family gatherings [1, 2].

Following a significant increase in diagnosed cases in late June 2020, the Victorian Government announced Stage 3 restrictions to apply from 9^th^ July 2020. All schools, cafes, restaurants and bars closed, and public gatherings and sporting events stopped, estimated to give a 60% community-wide contact reduction (CCR60) coupled with 100% school closure (SC100), as detailed in Table 1. Case isolation at home was assumed to have compliance of 70% for adults and 100% for children (CI70). The 20% workplace reduction continued, resulting in a (SC100+WN20+CCR60+CI70) Stage 3 social distancing strategy.

Seeding of infectious individuals into the model was stopped on 9^th^ July 2020, thus all further infections in the model occurred as a consequence of the breakdown in hotel quarantine measures and, subsequent large family gatherings. This replicates what is known to have occurred, with genomic sequencing recently showing that over 90% of all outbreak cases were due to a breakdown in quarantine measures, and arose from transmission between infected travellers and quarantine facility staff within a quarantine hotel [1, 2].

Calibrating our model to case data up to the beginning of August, and continuing simulations into 2021, allowed us to analyse the effectiveness of a range of alternative social distancing strategies, and to compare these with those applied by the Government of Victoria.

## Results

### Timing of response

The Government of Victoria adopted a “stepped” response to their second wave COVID-19 outbreak, from Stage 2 to Stage 3, then to Stage 4. This is illustrated by the yellow epidemic curve in Figure 2, which realistically predicts the dynamics of the reduction in daily case numbers up to 30^th^ October 2020, as seen in diagnosed case number data, Figure 1.

**Figure 2:**
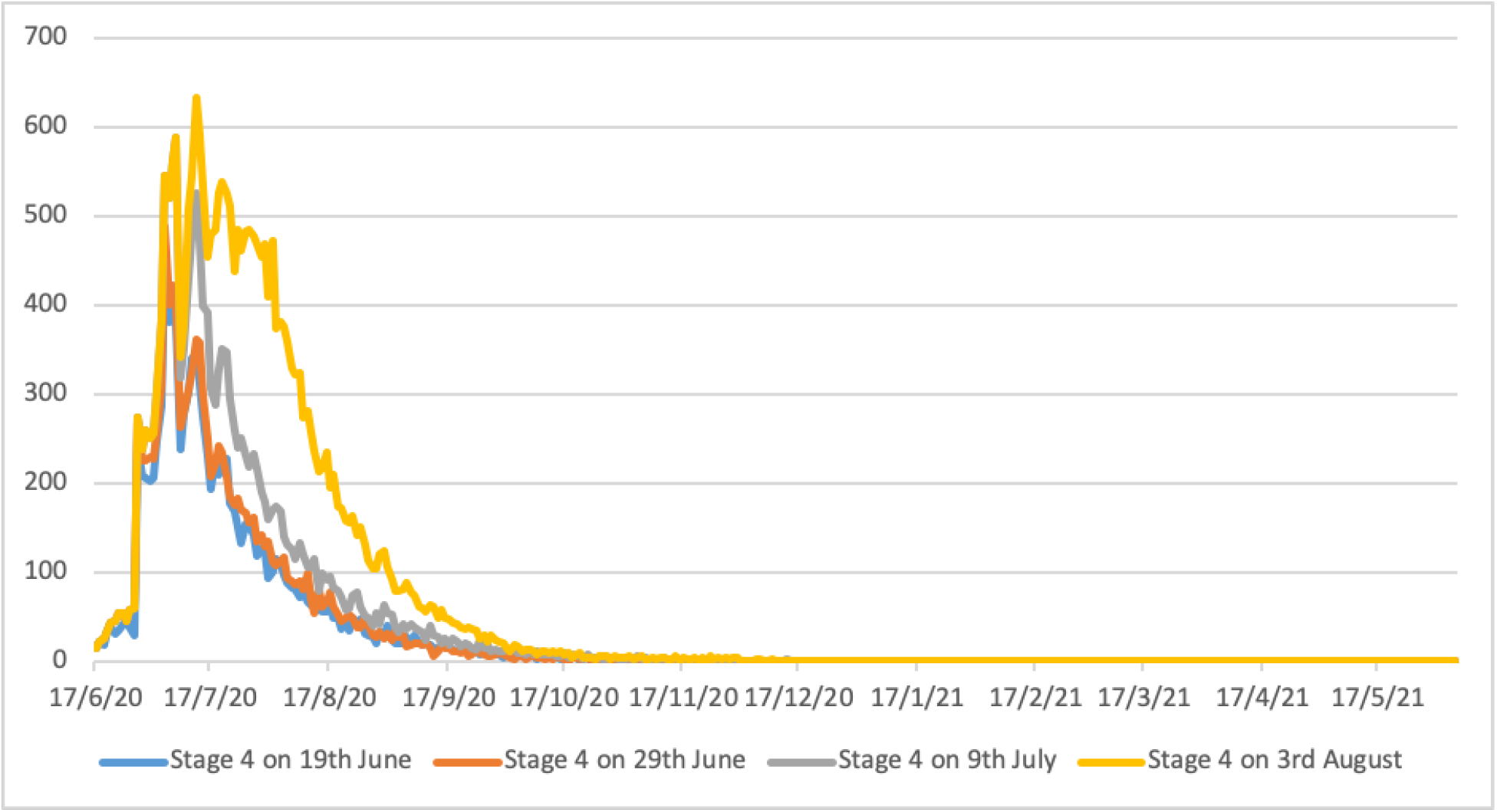
Predicted epidemic curves for earlier activation of Stage 4 lockdown measures in greater Melbourne (population 5 million). Y axis represents daily diagnosed cases. All scenarios assume schools are closed, and there is 90% case isolation for adults and 100% for children.

The rapid increase in diagnosed cases in greater Melbourne from 13^th^ June 2020 onwards (see Figure 1) suggests that an opportunity existed to move from Stage 2 measures directly to Stage 4. For reference, between 19^th^ June and 29^th^ June case numbers were doubling every 7 to 10 days. This exponential growth phase in diagnosed cases numbers may be observed in Figure 1. Figure 2, and data in Table 2, highlight the potential benefit of early activation of Stage 4 “lockdown” social distancing measures, on 19^th^ June, 29^th^ June, and 10^th^ July, the date when Stage 3 was activated, the blue, orange and grey curves respectively. These results illustrate the benefit, in terms of case number reductions, that may have been achieved by these early activation strategies, when compared with the actual date of Stage 4 lockdown on August 3^rd^, yellow curve Figure 2.

**Table 2:**
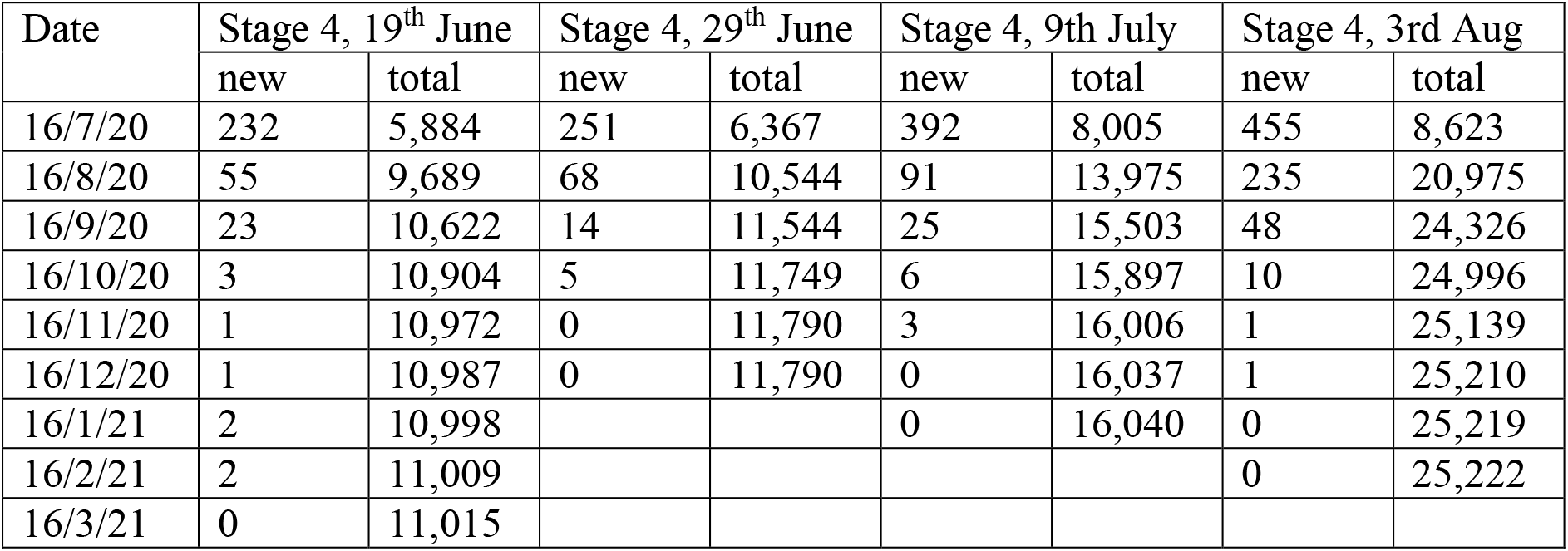
Predicted case numbers resulting from earlier Stage 4 activation in greater Melbourne (population 5 million). All scenarios assume schools are closed, and 70% case isolation for adults and 100% for children.

Table 2 highlights how moving to a Stage 4 “hard lockdown” on 9^th^ July would have reduced total case numbers by approximately 40%, from ∼25,000 to ∼16,000 cases, up to May 2020. These data correspond to the smaller area under the grey curve (10^th^ July activation) in Figure 2. When compared to the area under the yellow epidemic curve, which predict total cases resulting from the actual date of Stage 4 introduction on 3^rd^ August, this earlier activation is effective in reducing case numbers. Activating Stage 4 measures 10 to 20 days earlier, between 19^th^ and 29^th^ June, is shown to be highly effective, reducing total case numbers by over 50%.

Table 2 indicates that under all activation timings, Stage 4 measures are predicted to result in single digit case numbers by the end of October 2020, and effectively halt virus transmission by early 2021. Modelling the impact of the stepped measures introduced by the Government of Victoria, involving Stage 3 measures applied on 10^th^ July and Stage 4 on 3^rd^ August, accurately predicted the decline in case numbers. This is seen by comparing the yellow predicted 3^rd^ August epidemic curve in Figure 2, with the diagnosed case data up to 30^th^ October 2020, from Figure 1.

Throughout May 2020, daily case numbers in greater Melbourne had been steady and in single figures. As illustrated in Figure 1, reported daily case numbers began to increase throughout June and into July. Data generated by our modelling analyses (Table 2) predicts that Stage 4 lockdown activated on 29^th^ June, when daily case numbers had grown to 61 (see paragraph below), may have reduced total case numbers by over 50%. If activated 10 days later, on 9^th^ July, the reduction in total case numbers is less, reducing numbers by approximately 40%.

The following data sequence, taken from daily COVID-19 case data published by the Government of Victoria [7]; 5 cases on 11^th^ June, 10 on 13^th^, 20 on 16^th^, 25 on 19^th^, 33 on 24^rd^, 40 on 26^th^ and 61 on 29^th^ June 2020, 98 on 5^th^ July, 122 on 7^th^, 143 on 9^th^ and 218 on 14^th^ July [7, 8]. Note that reported case data is known to vary day-to-day, as seen in Figure 1, depending on numbers attending testing facilities. However, a clear pattern can be seen with this data sequence, with cases doubling approximately every 7-8 days from 11^th^ June to 9^th^ July, giving an exponential increase, then increasing linearly until mid-August, as illustrated in Figure 1 [8]. The above results suggest that the exponential increase in diagnosed cases during that period could have been used to trigger hard lockdown measures. If used, this triggering approach is predicted to effectively reduce virus transmission and resulting case numbers.

### Activation of stronger social distancing, 10th July 2020

The State of Victoria responded to the COVID-19 outbreak by increasing social distancing measures in two steps; Stage 3 measures on 10^th^ July 2020, and given the continuing increase in case numbers, strengthened measures to Stage 4 on 3^rd^ August 2020 [25]. Modelling was conducted to analyse the impact of this stepwise series of measures, and contrast these with the situation had they not occurred. Stage 3 measures introduced on 10^th^ July 2020, namely (SC100+WN20+CCR60+CI70), were strengthened to Stage 4 on the 3^rd^ of August, estimated to increase workplace non-attendance to 50% and further reduce community-wide contact to 20%, an 80% reduction. This gives social distancing scenario (SC100+WN50+CCR80+CI70), the grey curve in Figure 3.

**Figure 3:**
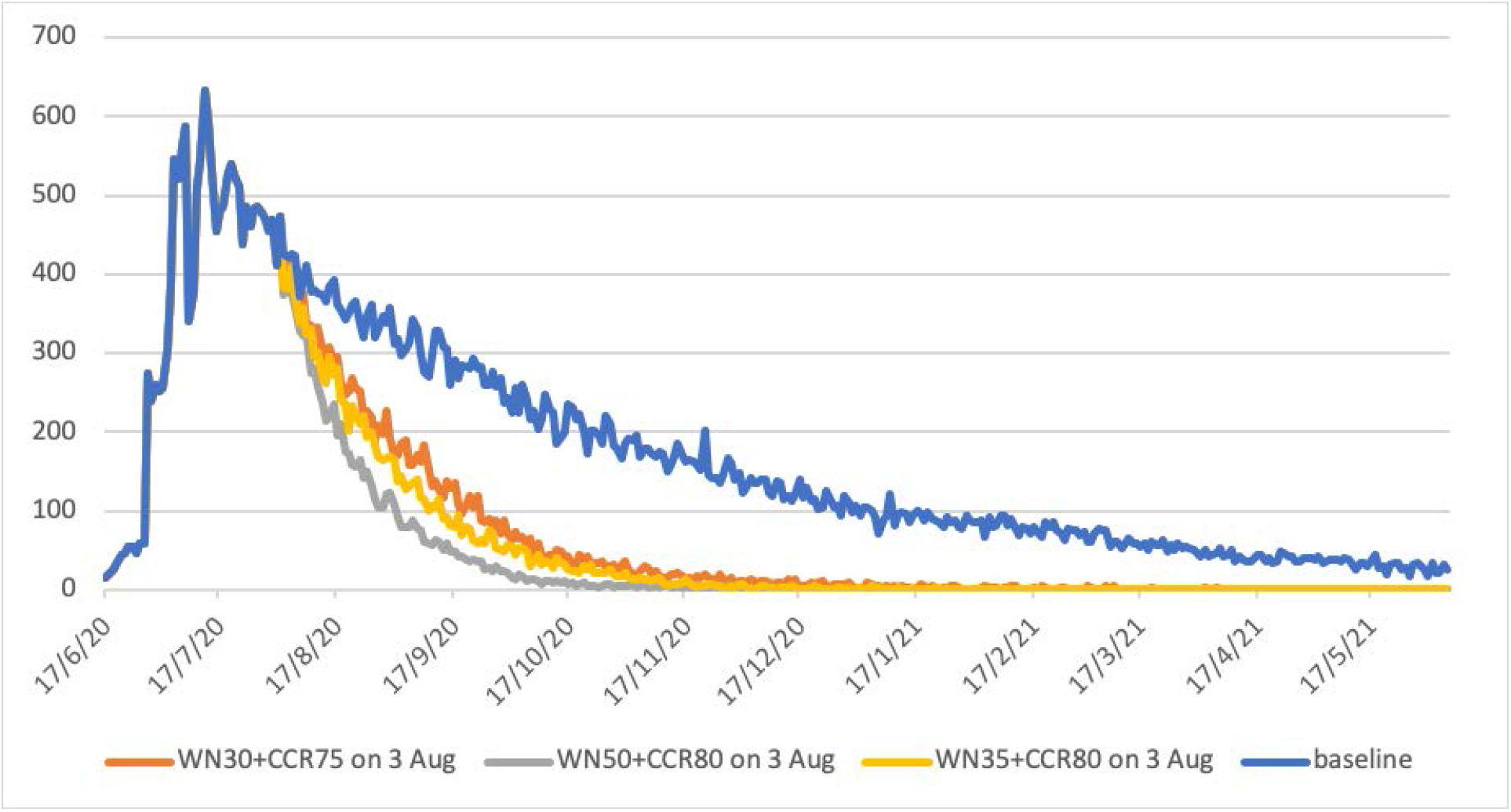
Increased social distancing activated on 3^rd^ August 2020 in greater Melbourne (population 5 million). Grey curve models Stage 4 measures applied. Yellow and orange curves allow greater workplace activity, increasing workplace attendance to 70% and 65% respectively. Blue curve represents continuation of Stage 3 measures, i.e. no increase in social distancing. All scenarios assume schools are closed, and there is 90% case isolation for adults and 100% for children.

Data generated by these modelling experiments (Table 3) highlight the need for, and effectiveness of, the hard lockdown measures taken, as illustrated by the grey curve in Figure 3, Without them, virus transmission is estimated to have continued to at least June 2021 had Stage 3 measures been maintained indefinitely, illustrated by the blue curve Figure 3.

**Table 3:** Increased social distancing measures activated on 3^rd^ August 2020, greater Melbourne (population 5 million). Daily cases mid-month and cumulative cases to 16^th^ May 2021. All scenarios have Stage 3 measures activated on 10^th^ July 2020, all schools are closed, and 70% case isolation for adults and 100% for children is assumed.

|  | Baseline Stage 3<br>10<sup>th</sup> July<br>(no Stage 4) |  | WN50 + CCR80<br>Stage 4<br>3<sup>rd</sup> August |  | WN30 + CCR75<br>3<sup>rd</sup> August |  | WN35 + CCR80<br>3<sup>rd</sup> August |  |
| --- | --- | --- | --- | --- | --- | --- | --- | --- |
|  | daily | total | daily | total | daily | total | daily | total |
| 16/08/20 | 393 | 22,313 | 235 | 20,975 | 288 | 21,642 | 271 | 21,387 |
| 16/09/20 | 260 | 32,393 | 48 | 24,326 | 129 | 27,628 | 81 | 26,365 |
| 16/10/20 | 200 | 39,801 | 10 | 24,996 | 42 | 29,883 | 32 | 27,960 |
| 16/11/20 | 170 | 45,630 | 1 | 25,139 | 17 | 30,735 | 9 | 28,488 |
| 16/12/20 | 121 | 49,875 | 1 | 25,210 | 14 | 31,082 | 2 | 28,601 |
| 16/01/21 | 93 | 53,088 | 0 | 25,219 | 0 | 31,230 | 0 | 28,643 |
| 16/02/21 | 72 | 55,725 | 0 | 25,222 | 1 | 31,306 | 0 | 28,650 |
| 16/03/21 | 59 | 57,658 |  |  | 1 | 31,355 |  |  |
| 16/04/21 | 41 | 59,161 |  |  | 0 | 31,382 |  |  |
| 16/05/21 | 29 | 60,294 |  |  | 0 | 31,386 |  |  |

Further modelling experiments were conducted to evaluate whether less restrictive Stage 4 strategies may have had a similar benefit, orange and yellow curves in Figure 3. The aim of these measures would be to lessen the economic impact of businesses closing due to hard lockdown, by increasing workplace attendance. This was achieved by *a)* allowing small service businesses to reopen, i.e. cafes with staff returning, and *b)* larger workplaces increasing numbers working in-situ, rather than remotely. We evaluated two scenarios: *1)* reducing workplace non-attendance to 30% (so increasing attendance to 70%) and lessening the community contact reduction to 75%, and *2)* having workplace non-attendance at 35% and community contact reduction staying at 80%. These result in the orange and yellow epidemic curves in Figure 5, respectively.

For all three lockdown strategies, these changes in social distancing are predicted to result in minimal virus transmission from mid-January 2021 onwards, with diagnosed case numbers at zero or one for all three strategies (see Table 3, Figure 3). The strategy reducing case numbers most rapidly corresponds to the Stage 4 measures that were adopted, and described previously (Figure 2, Table 2).

Strategies with a smaller reduction in workplace non-attendance, to (SC100+WN30+CCR75+CI70) or (SC100+WN35+CCR80+CI70), also result in case numbers decreasing rapidly. Unsurprisingly, the somewhat weaker alternative strategy of (SC100+WN30+CCR75+CI70) is slightly less effective in terms of outbreak duration, compared to that with workplace non-attendance at 35% and community contact reduction staying at 80%, Table 3. Overall, the reduction in the total number of cases is approximately linear according to the strength of measures, from 31,386 for the strategy with highest workplace attendance of 70% (WN30), to 28,650 for workplace attendance of 65% (WN35), and 25,222 for the Stage 4 measures used, with workplace attendance at 50% (WN50).

Predicted case data in Table 3 further indicate that all three increased social distancing strategies are effective in reducing epidemic duration, with daily case numbers in single figures by December 2020 or early 2021. This contrasts to the situation which would result if Stage 3 measures had remained in place, illustrated by the blue case curve in Figure 3, and case data in the first columns of Table 3. Under that scenario, virus transmission, and thus new cases, are predicted to occur up to and beyond May 2021.

### Early activation of Stage 3 measures

Additional modelling analyses were conducted to evaluate outcomes from earlier activation of Stage 3 measures. These had the aim of determining whether this change may have reduced the need for introduction of the more robust Stage 4 lockdown measures, and consequential negative economic impact. Figure 4 illustrate the impact of earlier activation of the Stage 3 social distancing measures, from (SC100+WN20+CCR60+CI70) to (SC100 +WN20+CCR60+CI70), five and ten days earlier that the actual 10^th^ July activation date.

**Figure 4:**
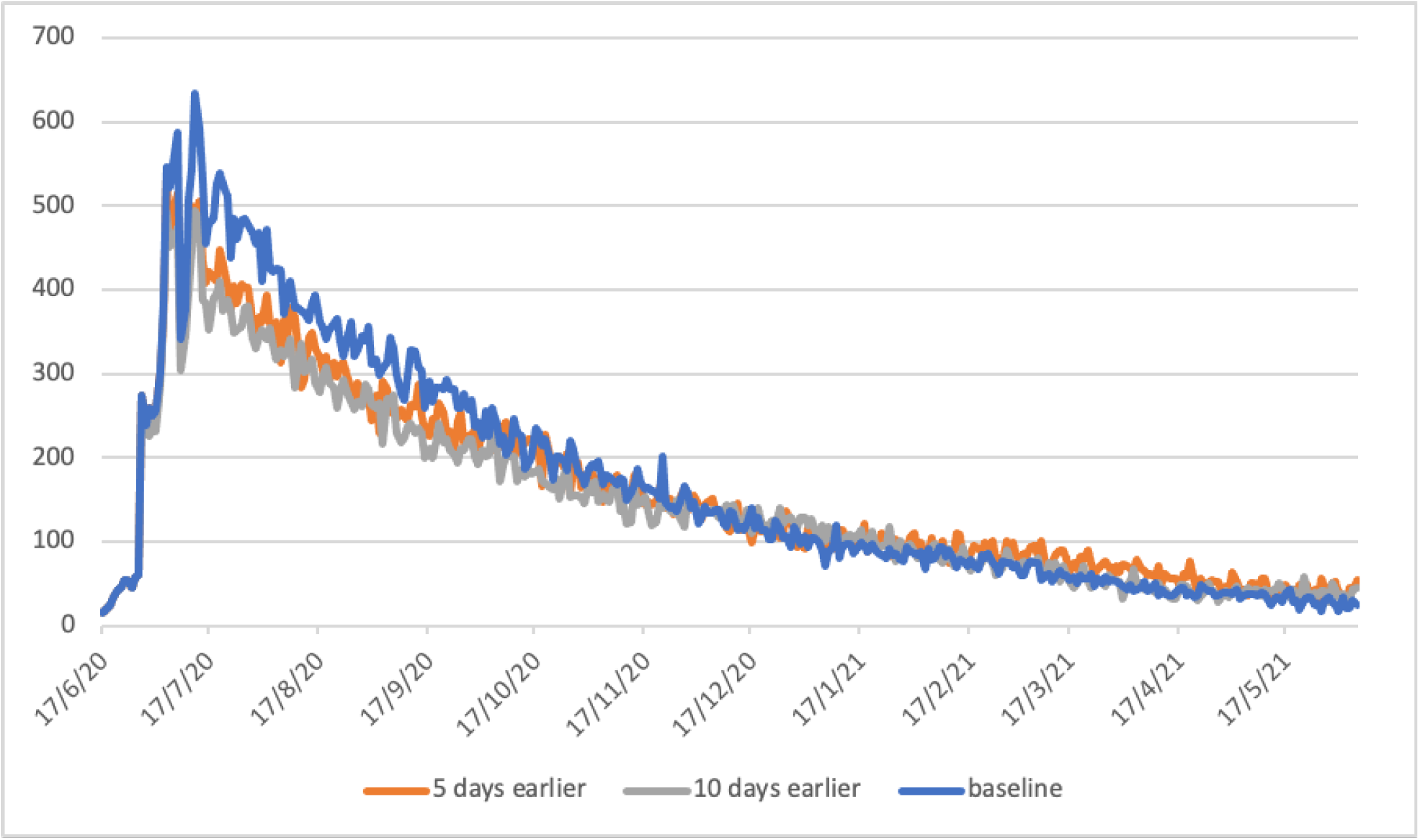
Greater Melbourne (population 5 million) COVID-19 outbreak with earlier Stage 3 social distancing activation. Blue curve, 9^th^ July 2020 actual date of activation; orange curve, earlier activation 4^th^ July; grey curve earlier activation on 29^th^ June.

Stage 3 activation on 29^th^ June, 10 days earlier than the 10^th^ July activation, are estimated to result in daily case numbers by mid-September reducing from 260 to 199. Total case numbers to mid-May 2021 are estimated to reduce from 60,294 to 52,475, Table 4. If activated 5 days earlier, on 4^th^ July, when daily case numbers were 64 [7], daily case numbers by mid-September drop from 260 to 233, and the total number up to mid-May 2021 will reduce to 56,920, Table 4.

**Table 4:**
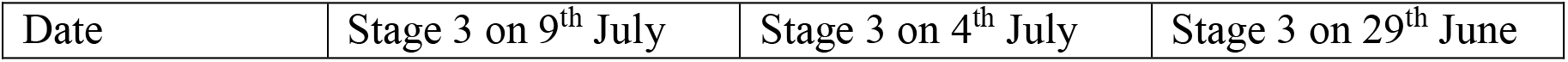
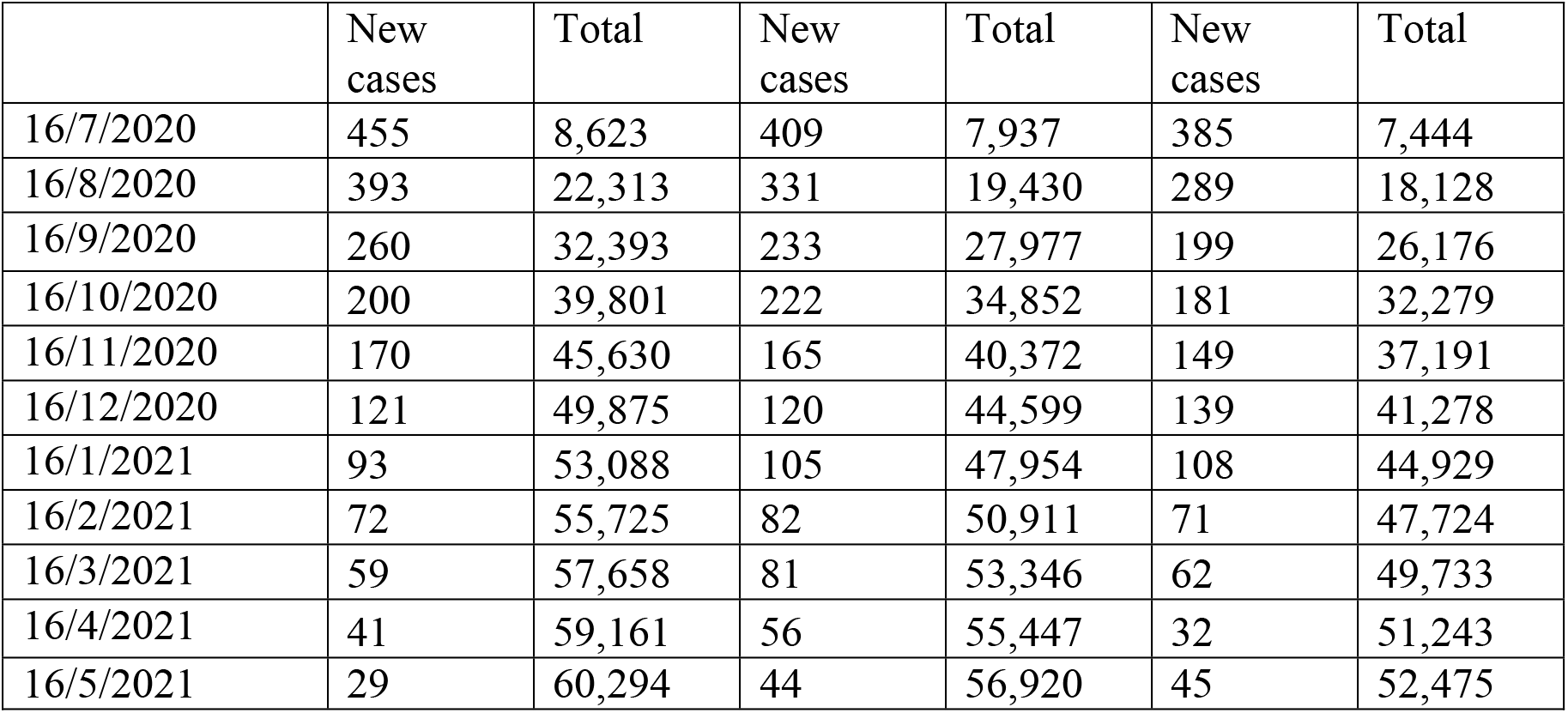
Daily and cumulative cases to 16^th^ May 2021 for greater Melbourne, population 5 million; Stage 3 social distancing activated on 9^th^ July (baseline), 4^th^ July and 29^th^ June 2020

| Date | Stage 3 on 9<sup>th</sup> July | Stage 3 on 4<sup>th</sup> July | Stage 3 on 29<sup>th</sup> June |
| --- | --- | --- | --- |

|  | New cases | Total | New cases | Total | New cases | Total |
| --- | --- | --- | --- | --- | --- | --- |
| 16/7/2020 | 455 | 8,623 | 409 | 7,937 | 385 | 7,444 |
| 16/8/2020 | 393 | 22,313 | 331 | 19,430 | 289 | 18,128 |
| 16/9/2020 | 260 | 32,393 | 233 | 27,977 | 199 | 26,176 |
| 16/10/2020 | 200 | 39,801 | 222 | 34,852 | 181 | 32,279 |
| 16/11/2020 | 170 | 45,630 | 165 | 40,372 | 149 | 37,191 |
| 16/12/2020 | 121 | 49,875 | 120 | 44,599 | 139 | 41,278 |
| 16/1/2021 | 93 | 53,088 | 105 | 47,954 | 108 | 44,929 |
| 16/2/2021 | 72 | 55,725 | 82 | 50,911 | 71 | 47,724 |
| 16/3/2021 | 59 | 57,658 | 81 | 53,346 | 62 | 49,733 |
| 16/4/2021 | 41 | 59,161 | 56 | 55,447 | 32 | 51,243 |
| 16/5/2021 | 29 | 60,294 | 44 | 56,920 | 45 | 52,475 |

Data in Table 4, and the epidemic curves in Figure 4, suggest that the 10 day earlier activation of Stage 3 measures on 29^th^ June has limited effect on both daily and overall case numbers, up to 17^th^ May 2021. The 5 day earlier activation of Stage 3 measures is predicted to reduce total case number by approximately 10%, while the 10 day earlier activation is predicted to reduce total case numbers by almost 20%.

These data indicate that the slightly earlier activation of Stage 3 social distancing measures fails to significantly reduce the duration of virus transmission, with transmission continuing to at least June 2021. This supports the decision by the Government of Victoria to then strengthen measures to Stage 4 lockdown, with our modelling estimating cessation of transmission by the end of 2020.

## Discussion

Early and robust application of social distancing measures are known to be an appropriate response to the COVID-19 pandemic. In early 2020 such responses were highly effective in minimizing case numbers and the rate of epidemic growth at the early stages of the pandemic, as in Korea and China [26-28]. Similarly, slow responses have resulted in significant first and second pandemic waves, *e*.*g*. in Italy, Spain and the UK [29].

This study’s aims were to provide further evidence on the benefit of, and need for, early and robust interventions to contain second waves of coronavirus infections. We quantified the effectiveness of a range of response measures, in terms of the reduction in case numbers, and thus resulting hospitalisations and deaths. These results are intended to help inform the hard COVID-19 containment decisions that need to be made by politicians and public health authorities, until an effective vaccine becomes available. A challenge in responding to COVID-19 second waves is to balance the effect of necessary lockdown measures, in protecting the health of the population, with the negative impact which strict social distancing measures have on a country’s economy. A fundamental issue highlighted in this study is the delay between increasing coronavirus transmission rates, and the time when these manifest themselves as an increase in diagnosed cases. Given that case data always lags the date of virus transmission, our findings indicate that activation of early lockdown is possibly the only feasible strategy to adopt.

Our study was fortunate in having access to comprehensive case data from the rapidly developing COVID-19 second wave in greater Melbourne, Australia from June 2020 onwards. Genomic sequencing has shown that most infections in this second wave result from a breakdown in hotel-based quarantine, followed by two large family gatherings [1, 2]. To our knowledge, there were no further introduced infections. This provided an ideal test-bed with which to analyse the effectiveness of the mitigation measures taken, determine their failings, understand why the second wave spread so rapidly, and to evaluate alternative mitigation strategies as to their effectiveness in reducing the scale of a second wave. These analyses were facilitated by the absence of introduced cases into the modelled population; the lack of “noise” from ongoing infection introduction allowed us to assume that all infections resulted from the single source.

Using second wave outbreak data, we demonstrate how the methodology used realistically modelled SARS-Cov-2 transmission from 1^st^ August to 30^th^ October 2020, when daily diagnosed case numbers had reduced to zero. This prediction of outbreak dynamics provides validation of how lockdown social distancing measures were modelled. That is, the impact which lockdown measures had on reducing transmission over the same period of time. This validation provides evidence as to the robustness of the modelling methods used, and gives credence to the results obtained.

Results indicate that the most effective response, which significantly reduce cases and second wave duration, would be the activation of Stage 4 lockdown measures much earlier, when case numbers were first seen to increase exponentially, on or before 29^th^ June 2020. This approach is estimated to result in ∼11,000 cases, compared to ∼25,000 for the strategy taken, in a population of approximately 5 million (Table 2). The second most effective strategy would have been to activate Stage 4 social distancing measures on 9^th^ July, the date of the first response to the second wave, which increased social distancing measures to Stage 3. This is estimated to reduce total cases to ∼16,000 (Table 2). These two highly effective strategies involved moving directly to Stage 4 measures from Stage 2, rather than the step-wise approach adopted, from Stage 2 to Stage 3 to Stage 4. They highlight the need to “catch” increasing transmission rates before infections are widely distributed, with early *and* robust social distancing contributing to the rapid reduction in virus transmission. The need for timely introduction of lockdown measures is discussed in a short overview of the situation in the UK at the end of October 2020, by Elisabeth Mahase [30]. It appears that the UK has been too slow to introduce hard lockdown, repeating the situation which occurred in greater Melbourne, but on a much larger, country-wide scale. Significantly, the easing of social distancing measures in the UK without high levels of testing and tracing has been shown in a recent modelling study to result in a second COVID-19 wave [31], predicting the situation which now appears to have occurred [30].

Had hard lockdown measures introduced on 3^rd^ August in Australia not occurred, we estimate virus transmission would be ongoing beyond the end of our simulation period (mid-May 2021), resulting in approximately 30 cases per day from February onwards (Figure 3, Table 3). While the Stage 3 social distancing measures activated in early July were predicted to results in a steady decrease in daily cases numbers, they did not lead to virus elimination by mid-2021. As a consequence, this would have limited the Government of Victoria’s ability to ease social distancing and prevent another COVID-19 wave. This would have been a similar situation to that analysed by Di Domenico and colleagues for Northern France [32, 33], and Aleta and colleagues for the North East of the USA [34].

Our analyses support the decision by the State Government to subsequently introduce Stage 4 lockdown measures, which rapidly suppressed ongoing transmission and shorten the duration of the second wave. Once transmission reaches a level where social distancing measures can be safely eased, and new cases managed by highly efficient test, track and isolate systems, this will permit an earlier reopening of businesses and a more rapid increase in economic activity.

It should be noted that the COVID-19 situation in Australia differs from most other countries, and while our key findings are widely applicable, they may not be as effective in other settings due to import of infectious cases. Prior to the second wave there was effectively no community transmission in Australia, due to early and strict border closures. Many countries in Europe, for example, failed to close borders, allowing infectious individuals to spread widely, with resulting widespread transmission, Secondly, the second wave of SARS-Cov-2 transmission was confined to the State of Victoria, particularly to the greater Melbourne area, and ongoing travel restrictions prevented introduction of further infections. This has resulted in the effective eradication of the virus, by early November 2020 [7].

Other countries experiencing second waves, or extensions of the first wave, are in a different situation, with limited ability to achieve virus elimination. Other means are needed to reduce transmission levels, and highly efficient testing, contact tracing and household isolation will be needed to contain case numbers to levels which prevent health care facilities from being overwhelmed. Modelling by Aleta and colleagues, in a USA setting [34], demonstrates how high levels of testing, with 50% of symptomatic cases diagnosed, 14 day quarantine of all members of a case’s household, and precautionary quarantine of contact households, may allow strict lockdown measures to be safely eased. They show that this level of testing and isolation is needed to prevent second COVID-19 waves. Similarly, modelling of methods to ease robust social distancing measures in the Paris region, also indicate that extensive testing and tracing are needed to manage second waves [32, 33]. The practical use of this response methodology can be seen with South Korea, which adopted a sophisticated test/trace/isolate approach from the early stages of the pandemic [26, 27]. The effectiveness of this approach has been shown to prevent second waves developing [35].

As with all model-based studies, there are limitations on what features we are able to replicate in detail, and what approximations need to be taken. These involve availability of data, both at the virus transmission level and the population level. We used an estimated basic reproduction number obtained from data gathered in Wuhan, China prior to social distancing activation, and used that to estimate the probability of transmission between two individuals. Detailed census data were used in model development, to create households, workplaces, and education establishments in as much detail as data sources permitted. This allowed us to model movement of individuals between their homes and work and education locations. However, mobility in the wider community was estimated based on the probability of random contact between pairs of individuals, weighted by distance from their homes. Obtaining data on actual population mobility, before and during periods of social distancing restrictions, would aid the fidelity of individual-based models such as ours, but obtaining such data is clearly a challenging task. While others have accessed de-identified mobile phone data to estimate movement throughout a population, as in[34], an example of how identified data can be accessed and applied in practice is given by South Korea. Here there is general support for government agencies having detailed location and mobility data for the whole population, as a public good. South Korea invested in Information Technology systems to manage future pandemics following the SARS outbreak, and has been highly successful in keeping COVID-19 case numbers low [26, 35]. Heavy use of mobile phone data to track the majority of the population, instructing individuals in geo-located “hotspot” areas to be tested, checking on compliance with home isolation, and informing residents of specific areas going into lockdown [26, 27, 35].

A further limitation of our study is the use of daily diagnosed case numbers as a surrogate for coronavirus transmission occurring between 5 to 15 days earlier, an approximate propagation time period from date of infection to symptom emergence, time to be tested, and the return of results. Diagnosed case data has a shorter propagation delay than daily hospitalisation and mortality data, and so gives an earlier indication of increasing levels of transmission, and can act as a trigger to increase social distancing measures, as described above. However, this use of case data suffers from the need to make assumptions on the percentage of the population who are asymptomatic following infection, and the percentage of symptomatic individuals who attend testing facilities. In this study we assumed that 35% of those infected were asymptomatic, and that approximately half of the remainder were diagnosed. COVID-19 hospitalisation data may act as a more accurate snapshot of previous rates of transmission, but with a longer propagation delay from time of infection.

The effect of social distancing measures on transmission rates was modelled directly, by reducing person-to-person contact in schools, workplaces and the wider community. These measures assumed that household contact remained. Case isolation was modelled by stopping movement of diagnosed cases out-with the home, allowing for a certain percentage of non-compliance. We assumed 70% compliance by adults and 100% for children, in the absence of published data sources. Availability of contact pattern survey data during the pandemic may have improved the fidelity of our model, and there is a hope that such data will be obtained and available for future pandemic situations.

### Health Policy Implications

Results from this study reinforce, and furthermore quantify, the benefit of early activation of robust response measures to second (and more) COVID-19 infection waves. Such measures are shown to significantly contain then reduce the epidemic growth rate, and consequential pressure on health care resources. Results demonstrate the criticality of the timing of activation, where a slow response to rapid, exponentially growing case numbers allows the coronavirus to spread widely within the population, before the introduction of more robust social distancing measures can take effect. The study also shows the benefit arising from border closure measures adopted for non-Australians by the Australian Government in late March 2020. These prevented the ongoing introduction of Sars-Cov-2 into the community, until a breakdown in international arrival quarantine measures initiated the second wave of transmission.

**I**n the absence of prospective testing, policy makers rely on diagnosed case, hospitalization and mortality data to inform decision making. All three metrics have inherent (and increasing) time lags; between date of infection and becoming infectious, and the possibility of being diagnosed, hospitalized or dying, with case data having the shortest “propagation delay”.

This study used a discrete COVID-19 second wave in Australia to demonstrate how exponentially increasing diagnosed case numbers, numbers that doubled every 7 to 10 days, could have better predicted the need for a significantly earlier activation of lockdown measures. Using daily case data as a lockdown “trigger” reinforces the need for a comprehensive and rapid testing program, as described in a related study of lockdown and exit strategies in France [10, 32]. As of early November 2020, many countries worldwide still lack highly effective testing and contact tracing systems, so limiting their ability to gain early warnings of a rapid growth in case numbers, and respond as indicated in this study.

A significant challenge for many countries facing second waves of COVID-19 cases is how to balance the negative effect which lockdown measures have on an economy, against the health of the population. It is clear there is a hesitancy by decision makers to introduce robust social distancing measures, as evidenced by the step-by-step approach adopted by the Government of Victoria, Australia, and evaluated in this study. This hesitancy is understandable. Long-duration school closure impacts education outcomes, particularly among those from low socio-economic backgrounds. Closure of cafes, restaurants and bars results in under-employment of young adults, and has a knock-on effect on the economy. Closure of service and transport industries results in increased unemployment, with these measures and home isolation impacting on the mental health of those affected.

Using data from the COVID-19 second wave in Australia, and analyzing alternative response strategies, the study determined the benefit of going hard and early. This contrasts with the approach take in Victoria, where social distancing measures were increased incrementally, resulting in the necessary hard lockdown measures being activated after the coronavirus was widely distributed within the population. This study suggests the optimal response strategy would be to go into lockdown much earlier, resulting in a significant reduction in case numbers, consequential hospitalisations and, potentially, a reduction in the mortality rate.

## Data Availability

Data contained in paper

## Supporting Information

**Table S1:**
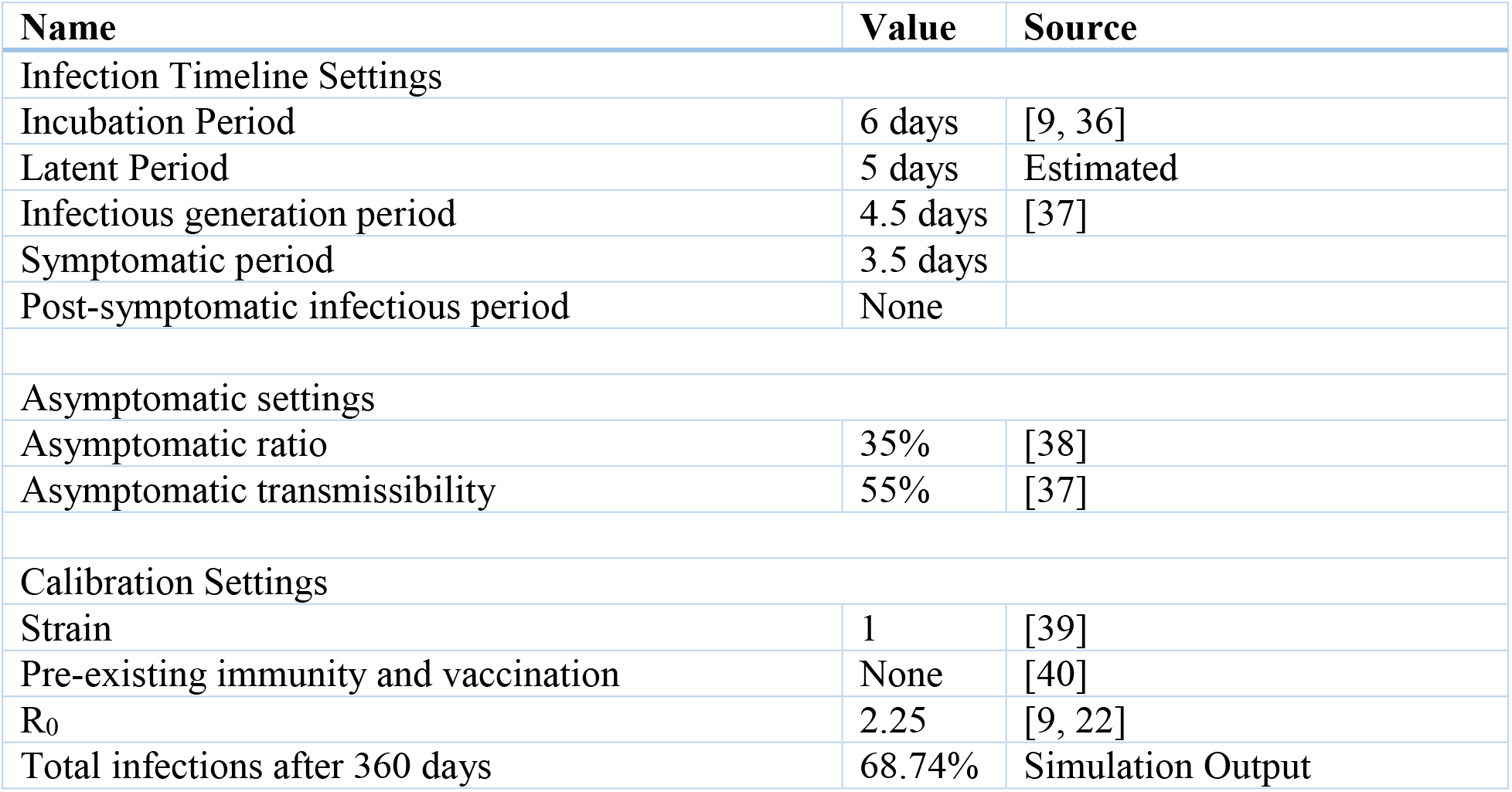
Summary of SARS-Cov-2 parameters

### Stage 3 and Stage 4 social distancing measures for greater Melbourne

Stage 3 restrictions (10^th^ July 2020)

> Stay at home restrictions, unless essential work, care-giving, shopping for essentials, medical needs.
>
> No travel from regional Victoria into metropolitan Melbourne.
>
> Restaurants and cafes takeaway only.
>
> Libraries and community venues closed except for hosting weddings, funerals, school use.
>
> Weddings limited to 5 people, funerals limited to 10 people.
>
> Cinemas, zoos, wildlife parks, galleries, museums, concert venues, campgrounds, caravan parks closed.
>
> Non-essential retail closed. Essential retail includes food/supermarkets, fuel, hairdressers etc.

Stage 4 restrictions, in addition to those for Stage 3 (3^rd^ August 2020)

> Schools closed
>
> Mandatory face masks outside home.
>
> Curfew in place from 8pm to 5am (must be at home, except for essential work, medical care, and caregiving).
>
> No visitors to home.
>
> Non-essential travel limited to 5km from home.
>
> Outdoor exercise limited to 1x 1hr session
>
> Only one person per household can leave home for necessary goods and services

